# Demographic Patterns, Social Determinants of Health, and Climate Change Perceptions Among Patients with Cancer and Hypertension

**DOI:** 10.64898/2026.01.21.26344565

**Authors:** Haoxin Chen, Jiancheng Ye

**Affiliations:** Weill Cornell Medicine, Cornell University, New York, NY, USA; Feinberg School of Medicine, Northwestern University, Chicago, IL, USA

**Author notes:** Corresponding author: Jiancheng Ye, PhD Weill Cornell Medicine, New York, NY, USA.

**Keywords:** cancer, hypertension, comorbidity, social determinants of health, health equity, climate change, health information

## Abstract

**Background:** The coexistence of cancer and hypertension presents complex clinical challenges, yet limited research has characterized this population using nationally representative data. Understanding demographic patterns, social determinants of health, and environmental health perceptions in this group is essential for developing targeted interventions.

**Methods:** We analyzed data from the Health Information National Trends Survey (HINTS) 6, focusing on 507 respondents with both cancer and hypertension (representing 12.82 million U.S. adults when weighted). We compared demographic, clinical, and psychosocial characteristics across sex and race/ethnicity groups using Chi-squared tests, Fisher’s exact tests, and Wilcoxon rank-sum tests. We examined responses to questions regarding social determinants of health, healthcare information sharing comfort, and climate change perceptions.

**Results:** Significant sex-based differences emerged, with females reporting lower marriage rates (49% vs. 75%, p < 0.001), lower income (33% vs. 17% earning <$35,000, p = 0.018), higher depression prevalence (31% vs. 18%, p = 0.043), and lower heart condition prevalence (16% vs. 31%, p = 0.012) compared to males. Racial/ethnic disparities were evident in diabetes prevalence (p = 0.012), with non-Hispanic White individuals showing the lowest rates (30%). Non-Hispanic Black and Asian respondents more frequently reported social determinant challenges including food insecurity, transportation barriers, and housing instability compared to non-Hispanic White respondents (all p < 0.05). Females perceived greater harm from climate change than males (50% vs. 41% responding “Some/A lot”, p = 0.013). Non-Hispanic Black respondents demonstrated the highest frequency of sun protection behaviors (90% reporting no recent sunburn).

**Conclusions:** Individuals with concurrent cancer and hypertension exhibit significant demographic and psychosocial heterogeneity. Sex-based and racial/ethnic disparities in social determinants of health, comorbidity patterns, and environmental health perceptions necessitate tailored, culturally responsive clinical interventions and policies to address the multifaceted needs of this population and reduce health inequities.

## INTRODUCTION

Cancer and hypertension represent two of the most prevalent chronic conditions posing significant challenges to global health systems. Cancer, characterized by the uncontrolled growth and spread of abnormal cells, remains a leading cause of morbidity and mortality worldwide. According to the World Health Organization (WHO), an estimated 10 million cancer-related deaths occurred in 2020.[1] Hypertension, or elevated blood pressure, affects approximately 1.13 billion individuals globally and serves as a major risk factor for cardiovascular disease, stroke, and kidney failure.[2] Both conditions impose substantial burdens on healthcare systems and profoundly impact the quality of life of affected individuals.[3]

The coexistence of cancer and hypertension in a single individual, termed comorbidity, presents a complex clinical scenario requiring integrated care approaches. Individuals with both conditions may experience unique challenges in healthcare management compared to those with either condition alone.[4] Certain chemotherapeutic agents, for instance, can exacerbate hypertension, leading to additional cardiovascular complications.[5] Understanding the demographic, clinical, and social characteristics of individuals with concurrent cancer and hypertension is crucial for optimizing their management and improving health outcomes.[6]

This study leverages data from the Health Information National Trends Survey (HINTS) 6, a nationally representative survey conducted by the National Cancer Institute (NCI) to assess the American public’s use of cancer-related information and health behaviors.[7] HINTS collects comprehensive data on topics related to cancer prevention, early detection, treatment, and survivorship, making it a valuable resource for studying the characteristics of individuals with cancer and hypertension.[8] By analyzing the HINTS 6 dataset, we aim to compare the demographic, clinical, and psychosocial characteristics of participants with concurrent cancer and hypertension to those without either condition. Additionally, we explore these characteristics stratified by sex and race/ethnicity, and examine associations with climate change perceptions and social determinants of health.[9]

While previous studies have explored the clinical characteristics of individuals with cancer or hypertension separately, few have comprehensively compared the characteristics of those with both conditions using nationally representative data. By identifying the unique features of this population, healthcare providers can develop tailored interventions to improve quality of life and reduce the burden of these chronic diseases. This paper contributes to understanding the intersectionality of cancer, hypertension, sex, race/ethnicity, climate change awareness, and social determinants of health, providing valuable insights for healthcare practitioners and policymakers to address health disparities and improve population health outcomes.

## METHODS

### Study population and data source

Data were obtained from the Health Information National Trends Survey (HINTS) 6, a nationally representative survey of the U.S. adult population.[7] The analysis focused on respondents reporting both cancer and hypertension diagnoses.

### Statistical analysis

Survey respondent demographic information was summarized and compared between sexes and among race/ethnicity groups within the subsample of HINTS respondents with both cancer and hypertension. Frequencies (%) were reported for categorical variables, while medians with interquartile ranges (IQR) were reported for continuous variables. Differences in these measures were evaluated using Chi-squared tests (or Fisher’s exact test for small sample sizes) and Wilcoxon rank-sum tests for categorical and continuous variables, respectively.

Differences in the frequency of survey responses were evaluated both between sexes and among race/ethnicity groups using Fisher’s exact test. These comparisons were also conducted across different medical diagnosis categories: no chronic conditions, cancer only, hypertension only, and both cancer and hypertension. All results are reported at a significance level of p < 0.05. Survey weights were applied to generate nationally representative estimates. All analyses were performed using R version 4.4.1.[10]

### Study approval

We solely used publicly available HINTS data for this study.

## RESULTS

### Sample characteristics

The analytic sample comprised 507 respondents with both cancer and hypertension, including 288 (56.8%) females and 219 (43.2%) males. When weighted to represent the general population, each female represented 7.12 million females and each male represented 5.70 million males in the U.S. population. The racial/ethnic composition included 370 (82%) non-Hispanic White, 72 (8.1%) non-Hispanic Black/African American, 45 (6.4%) Hispanic/Latino, 12 (2.0%) Asian, and 8 (1.9%) individuals reporting multiple race/ethnicities. Weighted to the population level, these groups represented: non-Hispanic White (10.46 million); non-Hispanic Black/African American (1.04 million); Hispanic/Latino (816,000); Asian (255,000); and individuals with multiple race/ethnicities (247,000).

Significant sex-based differences were observed in several demographic and clinical characteristics (**Table 1**). Compared to males, females demonstrated a lower frequency of being married (49% vs. 75%, p < 0.001), higher frequency of lower reported income (33% vs. 17%, p = 0.018), higher prevalence of depression (31% vs. 18%, p = 0.043), and lower prevalence of heart conditions (16% vs. 31%, p = 0.012).

**Table 1.** Demographic and clinical characteristics by biological sex among respondents with cancer and hypertension.

|  | **Female\***<br>**N = 288** | **Male\***<br>**N = 219** | **p-value\*\*** |
| --- | --- | --- | --- |
| **Race/Ethnicity** |  |  | 0.2 |
| Non-Hispanic White | 194 (78%) | 176 (86%) |  |
| Non-Hispanic Black | 54 (10%) | 18 (5.6%) |  |
| Hispanic/Latina | 27 (6.7%) | 18 (5.9%) |  |
| Asian | 7 (2.2%) | 5 (1.8%) |  |
| Other | 6 (3.0%) | 2 (0.6%) |  |
| **Age (y)** | 56 (46, 63) | 57 (50, 63) | 0.13 |
| **Occupation** |  |  | 0.12 |
| Employed | 68 (28%) | 54 (30%) |  |
| Unemployed | 12 (5.1%) | 6 (1.6%) |  |
| Retired | 166 (55%) | 154 (64%) |  |
| Other | 42 (13%) | 5 (4.2%) |  |
| **Marital status** |  |  | **<0.001** |
| Married | 98 (49%) | 143 (75%) |  |
| Not married | 188 (51%) | 76 (25%) |  |
| **Education** |  |  | 0.11 |
| HS or below | 81 (26%) | 43 (26%) |  |
| Some college/post-HS training | 101 (50%) | 65 (39%) |  |
| College graduate | 104 (25%) | 111 (35%) |  |
| **Sexual orientation** |  |  | 0.4 |
| Heterosexual | 269 (95%) | 207 (97%) |  |
| LGBTQ+ | 9 (4.7%) | 9 (2.5%) |  |
| Other | 3 (0.3%) | 1 (0.2%) |  |
| **Income ranges** |  |  | **0.018** |
| Less than \$35,000 | 108 (33%) | 45 (17%) | |
| \$35,000 to \$75,000 | 81 (32%) | 72 (33%) | |
| More than \$75,000 | 80 (35%) | 93 (50%) | |
| **General health** |  |  | 0.14 |
| Poor | 7 (1.2%) | 16 (5.4%) |  |
| Fair | 68 (17%) | 42 (16%) |  |
| Good | 117 (44%) | 88 (50%) |  |
| Very good | 81 (35%) | 60 (24%) |  |
| Excellent | 10 (3.6%) | 12 (4.6%) |  |
| **Medical conditions** |  |  |  |
| Diabetes | 99 (31%) | 83 (36%) | 0.3 |
| Heart condition | 51 (16%) | 67 (31%) | **0.012** |
| Lung disease | 66 (20%) | 38 (14%) | 0.3 |
| Depression | 86 (31%) | 42 (18%) | **0.043** |
| **Smoking status** |  |  | 0.4 |
| Current | 32 (11%) | 13 (6.2%) |  |
| Former | 97 (37%) | 98 (45%) |  |
| Never | 156 (52%) | 108 (49%) |  |
| **Neighborhood type** |  |  | 0.5 |
| Rural | 41 (17%) | 39 (20%) |  |
| Urban | 247 (83%) | 180 (80%) |  |
| **Census location** |  |  | 0.5 |
| South | 146 (50%) | 107 (44%) |  |
| West | 61 (19%) | 52 (19%) |  |
| Midwest | 50 (16%) | 29 (16%) |  |
| Northeast | 31 (14%) | 31 (21%) |  |

\* n (unweighted) (%)
\*\* Chi-squared test with Rao & Scott's second-order correction; Wilcoxon rank-sum test for complex survey samples

Significant differences emerged among race/ethnicity groups (**Table 2**). The non-Hispanic White population was relatively older (median age: 57 years) compared to other racial/ethnic groups (p = 0.015). Occupation differed significantly across groups (p = 0.037), with larger proportions of non-Hispanic White (61%), non-Hispanic Black (57%), and Hispanic/Latino (58%) populations being retired. Marital status varied significantly (p = 0.002), with a notably higher proportion of Asian respondents being married (98%). Educational attainment differed across groups (p = 0.003), with a higher proportion of Hispanic/Latino respondents (59%) having high school education or below. Regarding income (p = 0.024), non-Hispanic Black (44%) and Hispanic/Latino (41%) populations reported lower income ranges. The non-Hispanic White population demonstrated the lowest prevalence of diabetes (30%, p = 0.012). Neighborhood type differed significantly (p = 0.006), with individuals reporting multiple race/ethnicities more likely to reside in urban areas (60%).

**Table 2.** Demographic and clinical characteristics by race/ethnicity among respondents with cancer and hypertension.

|  | **Non-Hispanic White\***<br>**N = 370** | **Non-Hispanic Black\***<br>**N = 72** | **Hispanic/Latina\***<br>**N = 45** | **Asian\***<br>**N = 12** | **Other\***<br>**N = 8** | **p-value\*\*** |
| --- | --- | --- | --- | --- | --- | --- |
| **Sex** |  |  |  |  |  | 0.2 |
| Female | 194 (53%) | 54 (69%) | 27 (59%) | 7 (61%) | 6 (85%) |  |
| Male | 176 (47%) | 18 (31%) | 18 (41%) | 5 (39%) | 2 (15%) |  |
| **Age (y)** | 57 (49, 64) | 52 (41, 56) | 55 (42, 62) | 53 (47, 55) | 39 (36, 52) | **0.015** |
| **Occupation** |  |  |  |  |  | **0.037** |
| Employed | 83 (28%) | 18 (27%) | 12 (29%) | 5 (36%) | 4 (39%) |  |
| Unemployed | 10 (3.2%) | 3 (4.7%) | 5 (8.4%) | 0 (0%) | 0 (0%) |  |
| Retired | 245 (61%) | 42 (57%) | 26 (58%) | 4 (24%) | 3 (17%) |  |
| Other | 32 (7.5%) | 9 (12%) | 2 (4.7%) | 3 (40%) | 1 (44%) |  |
| **Marital status** |  |  |  |  |  | **0.002** |
| Married | 189 (64%) | 19 (38%) | 19 (41%) | 11 (98%) | 3 (68%) |  |
| Not married | 180 (36%) | 52 (62%) | 26 (59%) | 1 (2.3%) | 5 (32%) |  |
| **Education** |  |  |  |  |  | **0.003** |
| HS or below | 80 (23%) | 20 (33%) | 20 (59%) | 3 (20%) | 1 (3.2%) |  |
| Some college/post-HS training | 117 (47%) | 29 (45%) | 14 (28%) | 3 (27%) | 3 (36%) |  |
| College graduate | 171 (30%) | 23 (22%) | 11 (13%) | 6 (53%) | 4 (61%) |  |
| **Sexual orientation** |  |  |  |  |  | **<0.001** |
| Heterosexual | 350 (96%) | 63 (95%) | 45 (100%) | 10 (95%) | 8 (100%) |  |
| LGBTQ+ | 15 (4.3%) | 3 (3.2%) | 0 (0%) | 0 (0%) | 0 (0%) |  |
| Other | 1 (<0.1%) | 2 (1.5%) | 0 (0%) | 1 (5.0%) | 0 (0%) |  |
| **Income ranges** |  |  |  |  |  | **0.024** |
| Less than \$35,000 | 99 (24%) | 33 (44%) | 18 (41%) | 2 (7.1%) | 1 (3.2%) | |
| \$35,000 to \$75,000 | 105 (31%) | 24 (36%) | 16 (34%) | 5 (45%) | 3 (65%) | |
| More than \$75,000 | 145 (45%) | 13 (20%) | 7 (26%) | 4 (48%) | 4 (32%) | |
| **General health** |  |  |  |  |  | 0.3 |
| Poor | 18 (3.1%) | 3 (2.7%) | 2 (5.2%) | 0 (0%) | 0 (0%) |  |
| Fair | 71 (14%) | 23 (30%) | 11 (27%) | 2 (22%) | 3 (18%) |  |
| Good | 149 (48%) | 27 (40%) | 17 (28%) | 7 (52%) | 5 (82%) |  |
| Very good | 111 (31%) | 14 (24%) | 14 (40%) | 2 (18%) | 0 (0%) |  |
| Excellent | 18 (4.5%) | 3 (2.6%) | 0 (0%) | 1 (8.2%) | 0 (0%) |  |
| **Medical conditions** |  |  |  |  |  |  |
| Diabetes | 123 (30%) | 28 (48%) | 21 (35%) | 6 (45%) | 4 (77%) | **0.012** |
| Heart condition | 94 (24%) | 15 (19%) | 6 (19%) | 1 (2.3%) | 2 (15%) | 0.5 |
| Lung disease | 74 (16%) | 20 (28%) | 6 (16%) | 2 (14%) | 2 (49%) | 0.2 |
| Depression | 96 (24%) | 16 (33%) | 11 (17%) | 2 (11%) | 3 (60%) | 0.11 |
| **Smoking status** |  |  |  |  |  | 0.13 |
| Current | 29 (8.9%) | 12 (14%) | 3 (6.8%) | 1 (10%) | 0 (0%) |  |
| Former | 159 (44%) | 22 (34%) | 10 (20%) | 1 (12%) | 3 (17%) |  |
| Never | 180 (47%) | 38 (52%) | 31 (73%) | 10 (78%) | 5 (83%) |  |
| **Neighborhood type** |  |  |  |  |  | **0.006** |
| Rural | 69 (20%) | 6 (9.6%) | 2 (3.0%) | 0 (0%) | 3 (60%) |  |
| Urban | 301 (80%) | 66 (90%) | 43 (97%) | 12 (100%) | 5 (40%) |  |
| **Census location** |  |  |  |  |  | 0.2 |
| South | 176 (45%) | 44 (63%) | 25 (61%) | 5 (46%) | 3 (49%) |  |
| West | 84 (19%) | 5 (4.7%) | 15 (24%) | 6 (46%) | 3 (34%) |  |
| Midwest | 61 (18%) | 14 (15%) | 2 (2.3%) | 1 (8.2%) | 1 (11%) |  |
| Northeast | 49 (19%) | 9 (17%) | 3 (13%) | 0 (0%) | 1 (5.0%) |  |

\* n (unweighted) (%)
\*\* Chi-squared test with Rao & Scott's second-order correction; Wilcoxon rank-sum test for complex survey samples

In questions assessing social determinants of health, females (compared to males) more frequently responded “often/sometimes true” to household-level challenges (**Figure 1**). These included: cutting meal sizes or skipping meals due to insufficient money or food (HINTS question K1a; p < 0.001); inability to afford balanced meals (K1b; p < 0.001); worry about forced relocation (K1c; p < 0.001); and lack of reliable transportation affecting medical appointments, work, or daily activities (K1d; p < 0.001). Compared to non-Hispanic White respondents, non-Hispanic Black respondents responded “often/sometimes true” more frequently to questions K1a (p = 0.035), K1b (p = 0.022), K1c (p = 0.007), and K1d (p < 0.001) (**Figure 2**). Similarly, Asian respondents more frequently responded affirmatively to questions K1a (p = 0.030), K1b (p = 0.013), and K1d (p = 0.038) compared to non-Hispanic White respondents. No significant differences were observed between non-Hispanic White and Hispanic/Latino respondents or other race/ethnicities. Compared to respondents with both cancer and hypertension, those with hypertension alone responded “often/sometimes true” more frequently to questions K1a (p < 0.001), K1b (p = 0.001), and K1d (p = 0.010) (**Figure 3**). Respondents with cancer only more frequently endorsed K1b (p = 0.018). Similarly, those without cancer or hypertension responded affirmatively more frequently to question K1a (p = 0.018) compared to the cancer and hypertension group.

**Figure 1.**
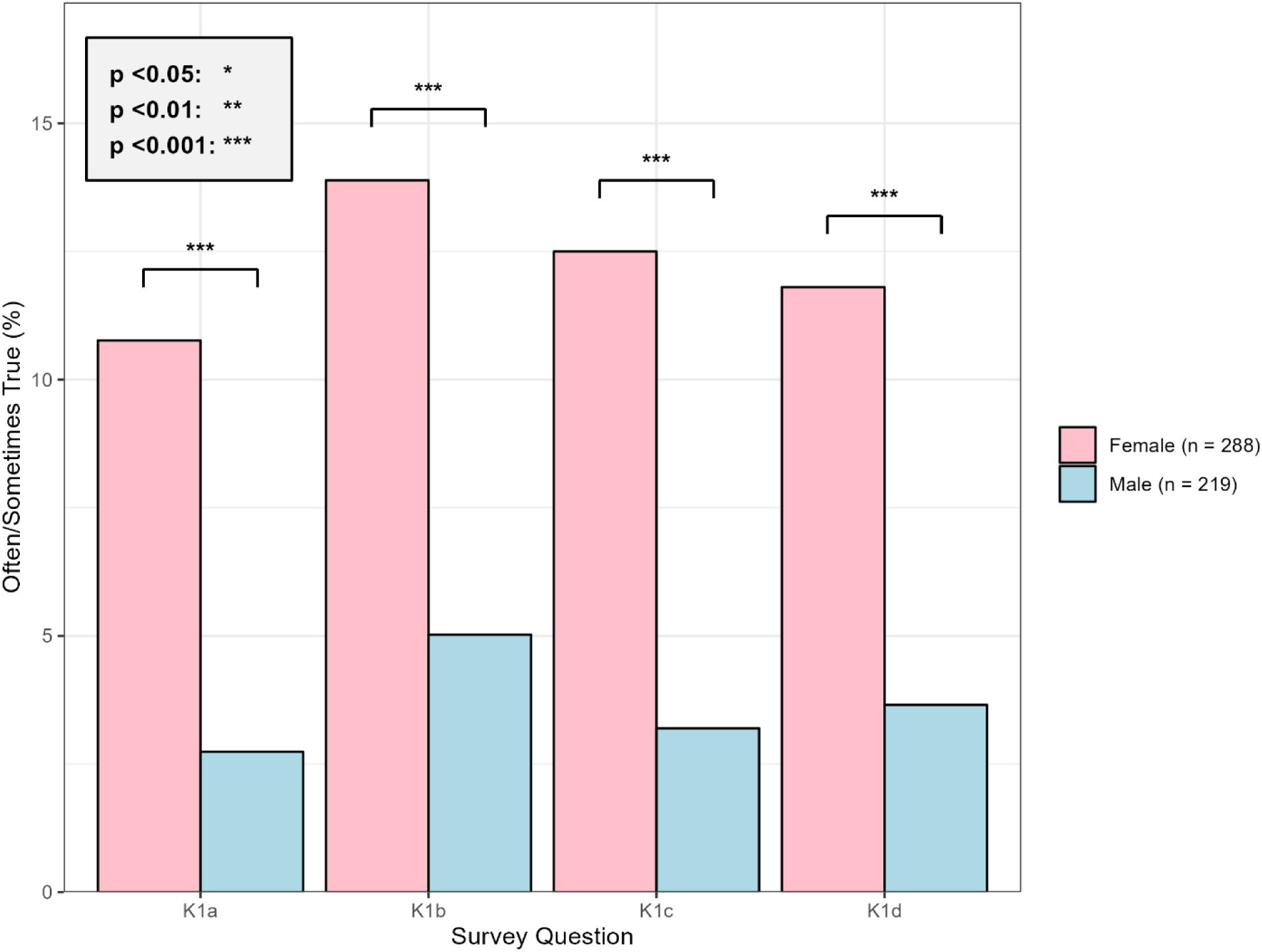
Social determinants of health challenges by biological sex among respondents with cancer and hypertension. Bar chart showing the percentage of male and female respondents who answered “often/sometimes true” to questions regarding household food insecurity (K1a: cutting meal sizes or skipping meals; K1b: inability to afford balanced meals), housing instability (K1c: worry about forced relocation), and transportation barriers (K1d: lack of reliable transportation affecting medical appointments, work, or daily activities). Error bars represent 95% confidence intervals.

**Figure 2.**
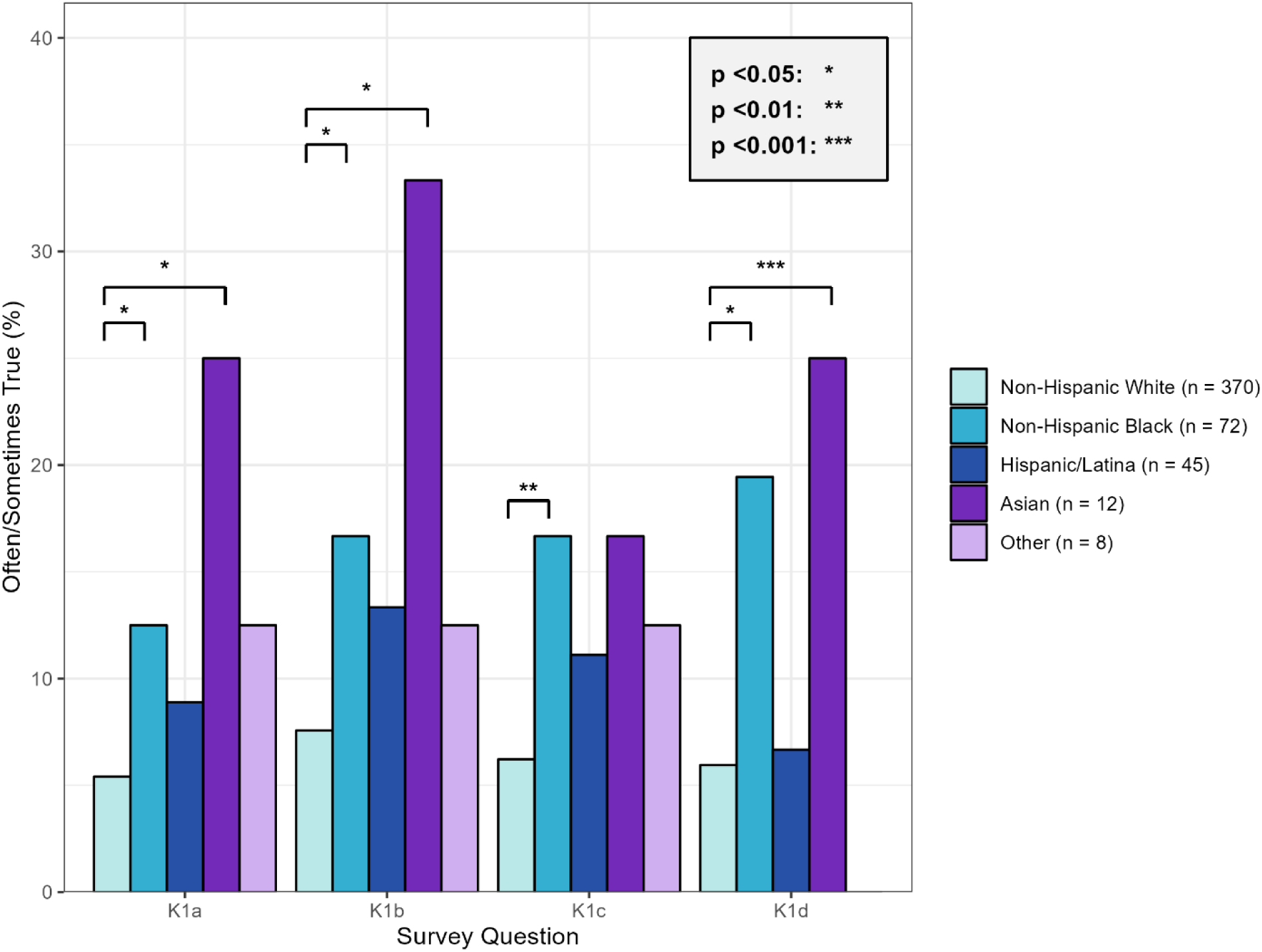
Social determinants of health challenges by race/ethnicity among respondents with cancer and hypertension. Bar chart displaying the percentage of respondents across different racial/ethnic groups (non-Hispanic White, non-Hispanic Black, Hispanic/Latino, Asian, and Other) who answered “often/sometimes true” to questions regarding food insecurity (K1a, K1b), housing instability (K1c), and transportation barriers (K1d). Error bars represent 95% confidence intervals. Asterisks indicate statistical significance compared to non-Hispanic White respondents.

**Figure 3.**
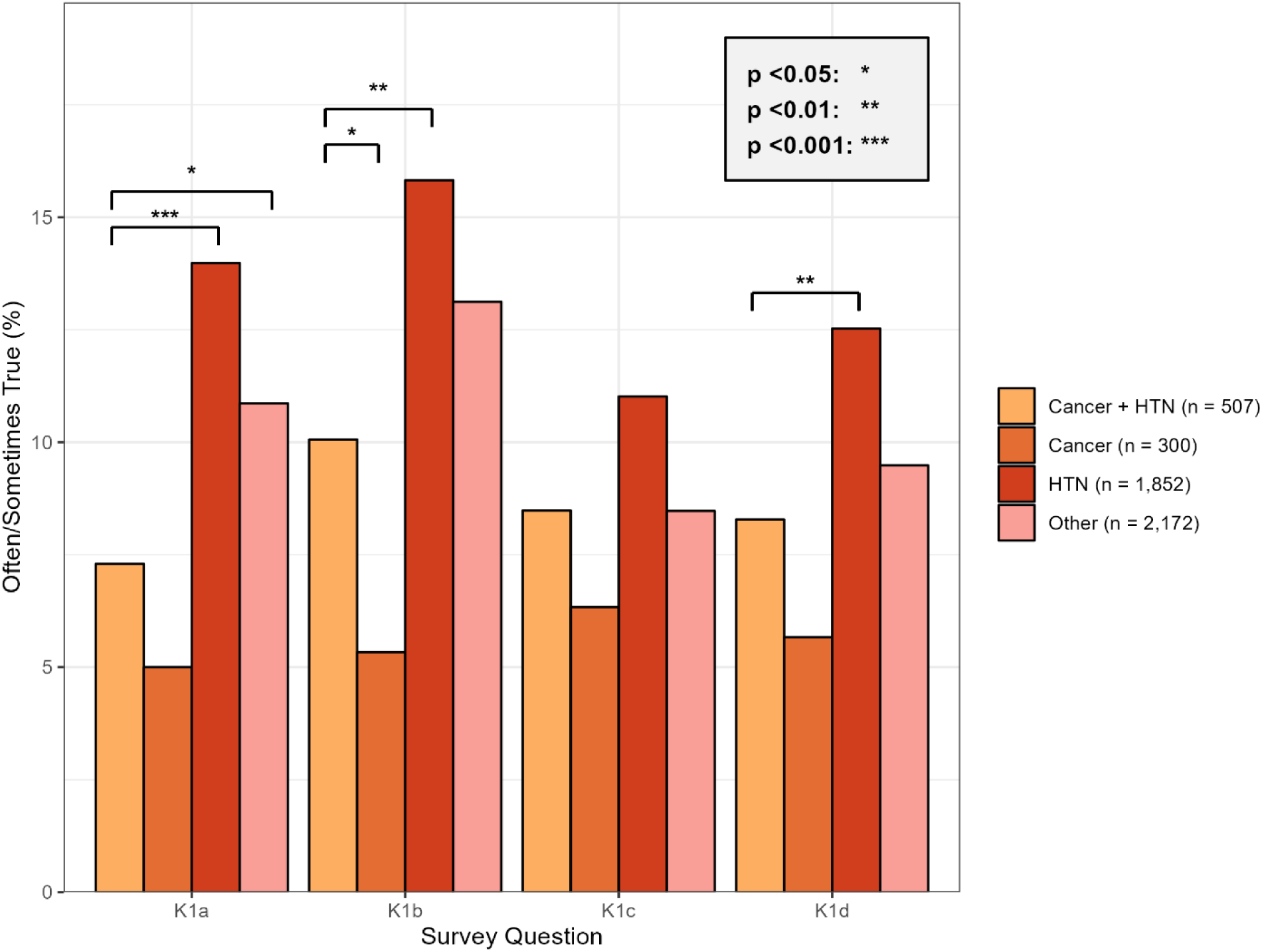
Social determinants of health challenges by disease status. Bar chart comparing the percentage of respondents who answered “often/sometimes true” to social determinant questions (K1a-K1d) across four groups: no chronic conditions, cancer only, hypertension only, and both cancer and hypertension. Error bars represent 95% confidence intervals. Asterisks indicate statistical significance compared to the cancer and hypertension group.

No significant differences were observed between males and females regarding comfort with healthcare providers sharing information for treatment purposes related to: affording or accessing healthy food (HINTS question K2a); transportation difficulties (K2b); or housing issues (K2c) (**Figure 4**). Compared to non-Hispanic White respondents, non-Hispanic Black respondents more frequently responded “very/somewhat comfortable” to questions K2b (p = 0.041) and K2c (p = 0.029) (**Figure 5**). Compared to respondents with cancer and hypertension, those with hypertension alone less frequently indicated comfort sharing information for questions K2b (p = 0.004) and K2c (p = 0.018). Similarly, those without cancer or hypertension expressed lower comfort levels for questions K2a (p = 0.002), K2b (p = 0.005), and K2c (p = 0.001) (**Figure 6**).

**Figure 4.**
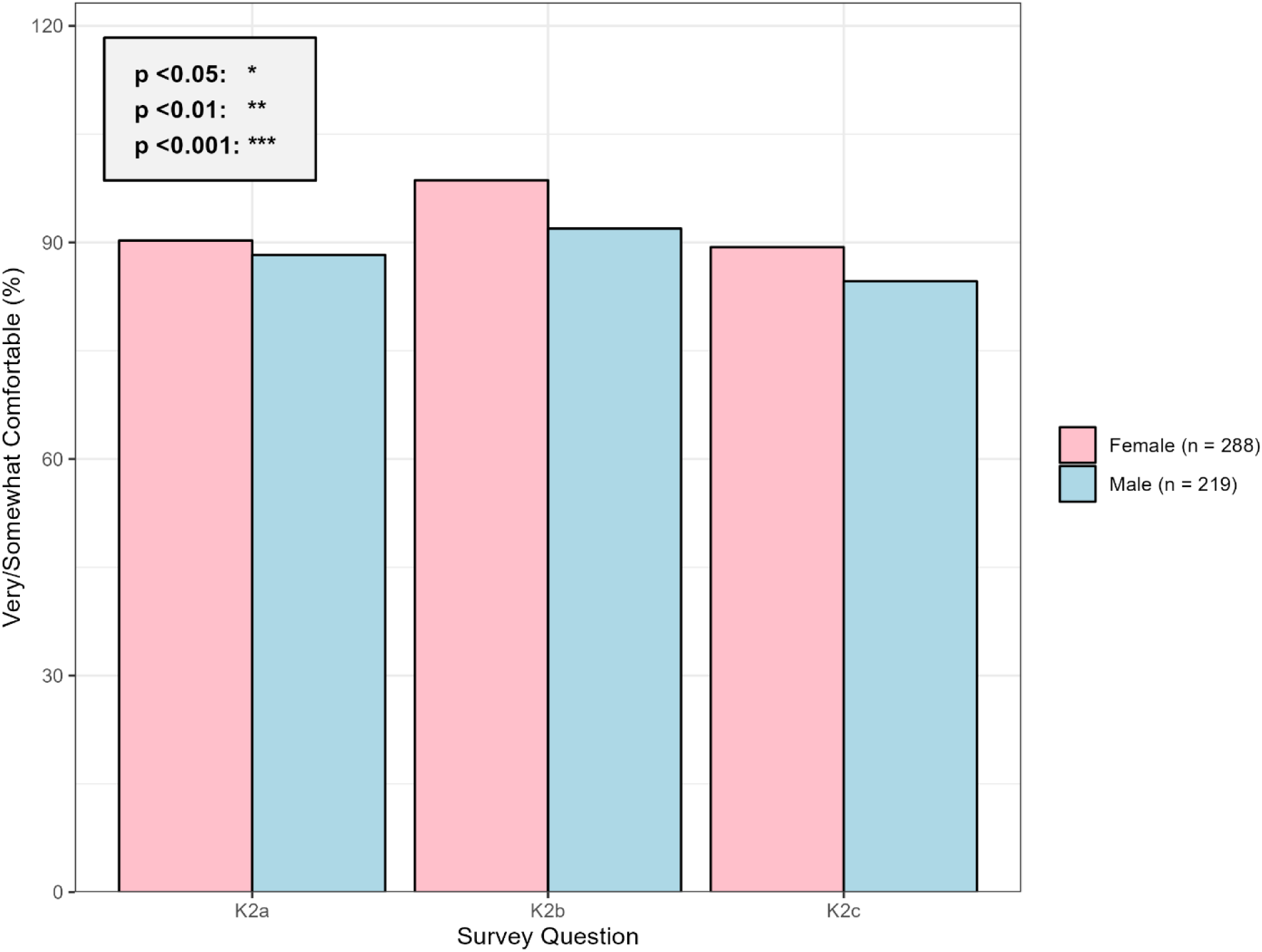
Comfort with healthcare information sharing by biological sex among respondents with cancer and hypertension. Bar chart showing the percentage of male and female respondents who indicated they would be “very/somewhat comfortable” with healthcare providers sharing their information for treatment purposes related to food access (K2a), transportation difficulties (K2b), and housing issues (K2c). Error bars represent 95% confidence intervals. No significant differences were observed between sexes.

**Figure 5.**
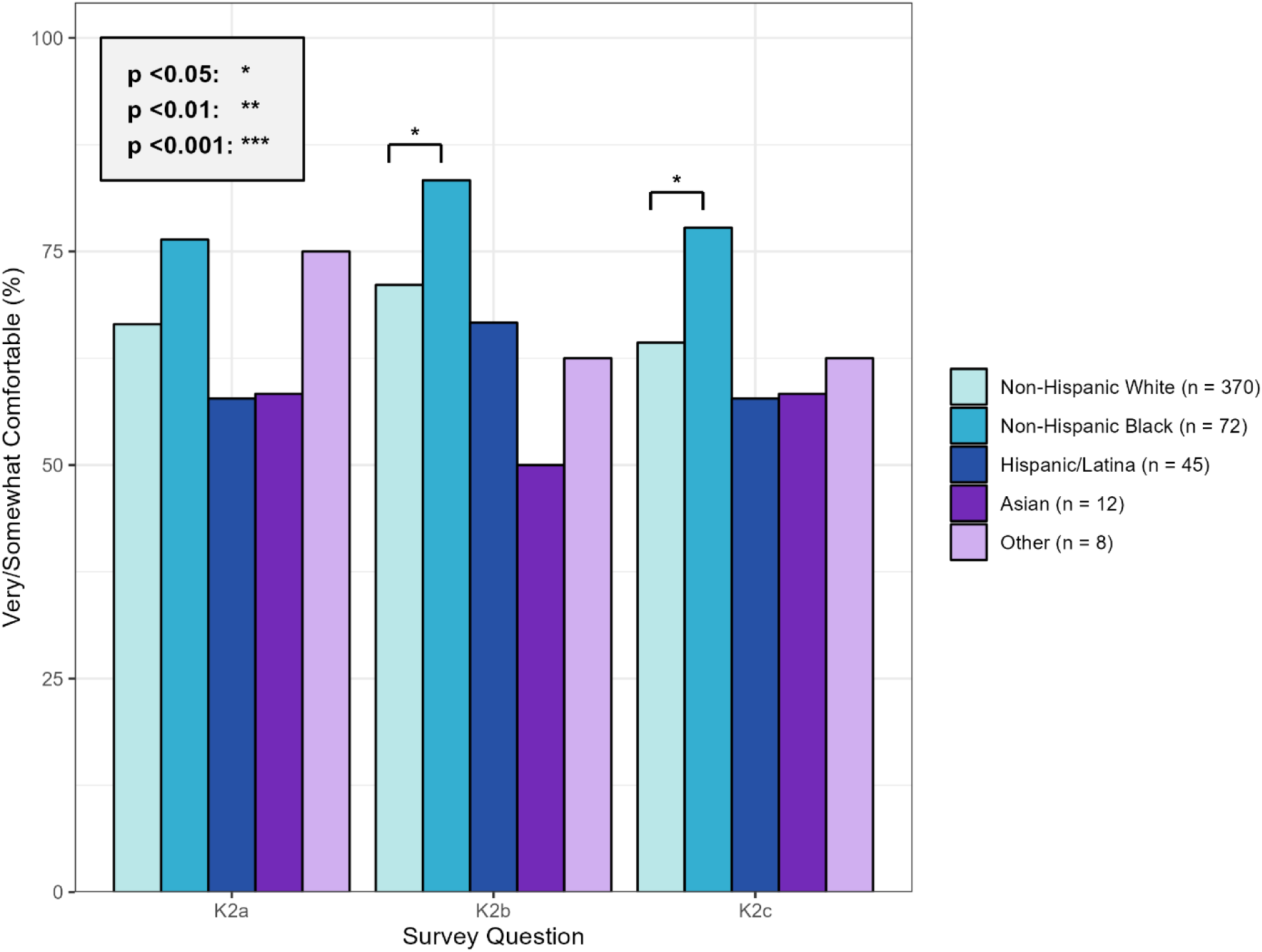
Comfort with healthcare information sharing by race/ethnicity among respondents with cancer and hypertension. Bar chart displaying the percentage of respondents across different racial/ethnic groups who indicated they would be “very/somewhat comfortable” with healthcare providers sharing their information for treatment purposes related to food access (K2a), transportation difficulties (K2b), and housing issues (K2c). Error bars represent 95% confidence intervals. Asterisks indicate statistical significance compared to non-Hispanic White respondents.

**Figure 6.**
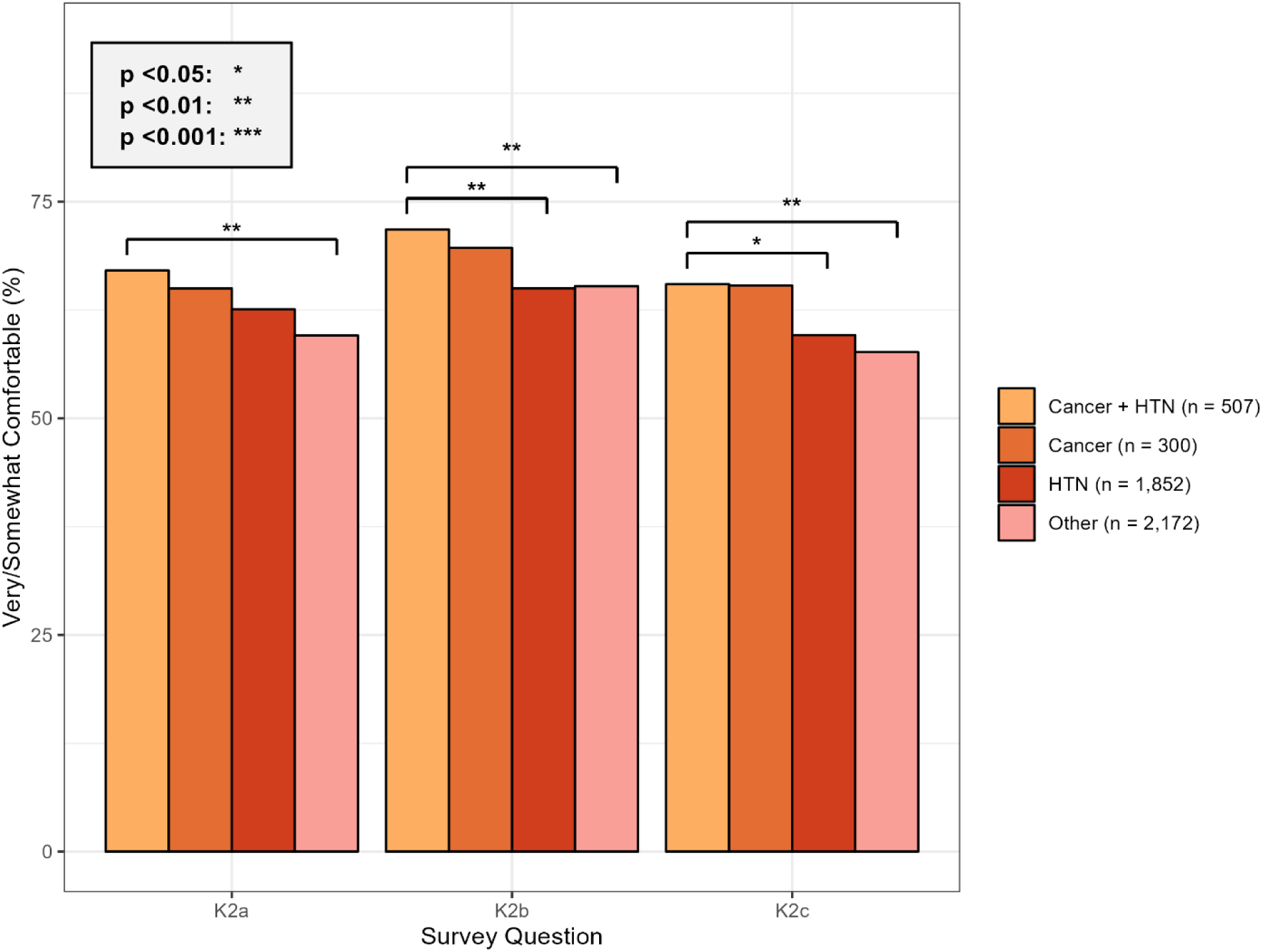
Comfort with healthcare information sharing by disease status. Bar chart comparing the percentage of respondents who indicated they would be “very/somewhat comfortable” with healthcare information sharing (questions K2a-K2c) across four groups: no chronic conditions, cancer only, hypertension only, and both cancer and hypertension. Error bars represent 95% confidence intervals. Asterisks indicate statistical significance compared to the cancer and hypertension group.

Figure 7 presents a sankey diagram illustrating the relationships among sex, race/ethnicity, and environmental health perceptions. When asked about activities during their most recent sunburn, responses categorized as “One activity” included: working outside at home or at a family/friend’s home (n = 22), sunbathing (n = 9), exercise (n = 8), attending outdoor events (n = 4), day-to-day activities (n = 4), swimming (n = 4), watching sporting events (n = 4), and working at an outdoor job (n = 2). Regarding perceived harm from climate change, females more frequently responded “Some/A lot” (50% vs. 41%) and “Don’t know” (18% vs. 13%), while less frequently responding “A little/Not at all” (32% vs. 46%) compared to males (p = 0.013). No significant differences were observed among race/ethnicity groups for this question. When asked about activities during their most recent sunburn, females were more likely to respond “No sunburn” (80% vs. 69%) compared to males (p = 0.006). Significant differences emerged among race/ethnicity groups, with non-Hispanic Black respondents demonstrating the highest frequency of “No sunburn” responses (90%) compared to non-Hispanic White (72%), Hispanic/Latino (80%), and Asian respondents (80%). Respondents reporting “No sunburn” demonstrated different response patterns when asked about climate change harm (p < 0.001). Specifically, “No sunburn” respondents more frequently responded “Don’t know” (17% vs. 11%) and less frequently “A little/not a lot” (38% vs. 41%) and “Some/A lot” (45% vs. 49%).

**Figure 7.**
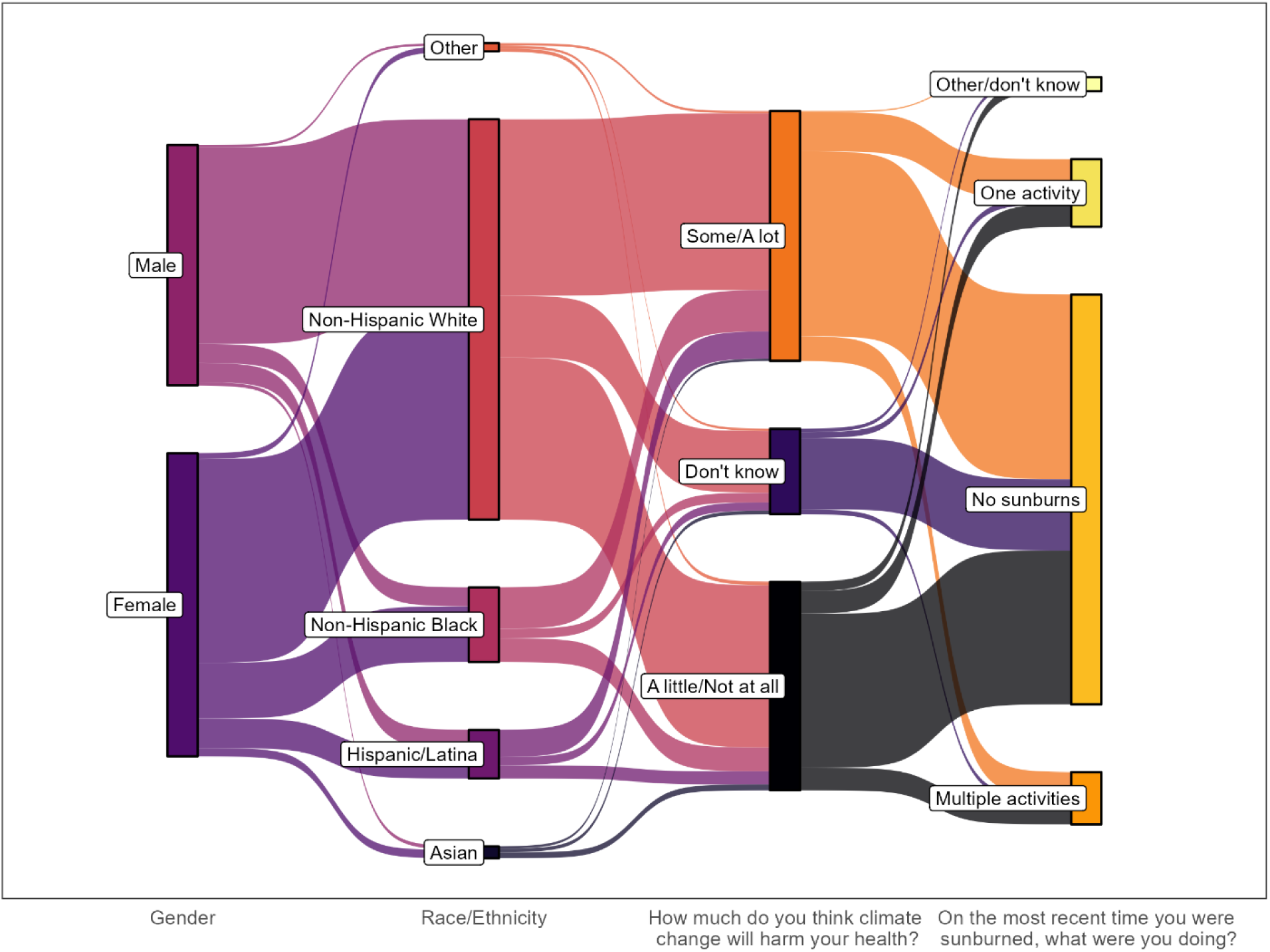
Sankey diagram illustrating relationships among biological sex, race/ethnicity, and environmental health perceptions. The diagram shows the flow and connections between demographic characteristics (sex and race/ethnicity) and responses to questions about sun exposure (”No sunburn” vs. “One activity”) and perceived harm from climate change (”Don’t know,” “A little/Not at all,” “Some/A lot”). Line thickness represents the proportion of respondents in each category. The visualization demonstrates how demographic factors relate to environmental health awareness and behaviors among individuals with cancer and hypertension.

## DISCUSSION

This study provides valuable insights into the management needs of patients with cancer and hypertension, revealing significant patterns across demographics, health status, social determinants, and climate change perceptions. We identified notable sex-based differences in social challenges and healthcare information sharing preferences, as well as racial/ethnic disparities in social determinants of health. Additionally, patients with different chronic condition profiles face varying levels of social and economic challenges. Variations in climate change perceptions across demographic groups underscore the need for targeted educational efforts. These findings suggest that tailored interventions and policies are necessary to address diverse patient needs and improve health outcomes across populations.[11]

### Targeted treatment for comorbidities

Our analysis revealed distinct patterns in comorbid conditions across sexes, suggesting the need for sex-specific clinical interventions. Women demonstrated higher rates of depression, indicating the need for integrated mental health screening and treatment programs that include counseling, peer support groups, and evidence-based stress management interventions.[12, 13] For men, who showed higher rates of cardiovascular disease, interventions should emphasize cardiovascular risk reduction through regular screening, dietary modifications, structured exercise programs, and medication adherence support.[14, 15]

The observed racial/ethnic disparities in chronic disease prevalence highlight the need for culturally tailored lifestyle interventions. Healthcare systems should provide culturally appropriate resources to promote healthier lifestyles, including personalized dietary counseling reflecting cultural food preferences, community-based physical activity programs, and health education workshops delivered in culturally sensitive formats. The higher prevalence of diabetes among non-Hispanic Black, Asian, and other minority groups necessitates enhanced disease management approaches, including intensified screening programs, culturally adapted diabetes self-management education, and targeted support services addressing specific risk factors and common comorbidities in these communities.[16] Implementing these evidence-based, culturally responsive strategies can help reduce health disparities and improve clinical outcomes across demographic groups.

### Addressing social determinants of health

Our study revealed significant sex-based differences, racial/ethnic disparities, and health condition-specific challenges related to social determinants of health. Females more frequently reported food insecurity, transportation barriers, and housing instability compared to males. Non-Hispanic White respondents less frequently endorsed social determinant challenges compared to non-Hispanic Black and Asian respondents, suggesting differential vulnerability or reporting patterns. Notably, patients with hypertension alone faced more pronounced social determinant challenges than those with cancer alone or both conditions.

Integrating social determinants of health into clinical care strategies is essential for improving patient outcomes.[17] For women, programs should prioritize access to nutritional assistance, reliable transportation services, and affordable housing resources. Understanding and addressing racial/ethnic disparities requires systematic efforts to ensure equitable support across all communities.[18] Policy initiatives should focus on identifying root causes of these disparities and implementing targeted interventions informed by community engagement.[19] For patients with hypertension, community-based programs offering comprehensive education, resource navigation, and peer support for hypertension management may be particularly beneficial.[20]

Our findings also revealed differential comfort levels with healthcare information sharing across demographic groups. Females demonstrated greater comfort with information sharing among providers, potentially reflecting higher trust in healthcare systems or greater recognition of the value of care coordination. Conversely, non-Hispanic White respondents expressed lower comfort compared to non-Hispanic Black respondents, suggesting potential trust differentials that warrant further investigation. Patients with concurrent cancer and hypertension exhibited higher comfort levels with information sharing, likely reflecting their greater need for coordinated, integrated care across multiple providers.[21]

To address these patterns, healthcare systems should enhance transparency and communication to improve patient comfort and outcomes, particularly for women who appear more receptive to collaborative care models. Investigating the underlying factors contributing to lower comfort among non-Hispanic White individuals could inform strategies to build trust equitably across racial/ethnic groups. Promoting integrated care models that facilitate seamless, secure information sharing among providers is crucial for patients with complex health needs, particularly those managing multiple chronic conditions.[22] These patient-centered policies can significantly enhance care coordination and overall health outcomes.

### Climate change perceptions and health behaviors

Our research revealed significant insights into climate change perceptions and sun exposure behaviors across demographic groups. Women perceived greater potential harm from climate change and expressed more uncertainty compared to men, suggesting heightened awareness or concern regarding environmental health impacts. This pattern indicates opportunities for targeted health communication that acknowledges these concerns while providing evidence-based, actionable information to reduce climate-related health risks and promote adaptive behaviors.[23]

Notably, we observed no significant differences in climate change harm perceptions across racial/ethnic groups, underscoring broad recognition of climate change as a public health concern that transcends demographic boundaries. The substantially higher frequency of “no sunburn” responses among non-Hispanic Black respondents reflects varying sun exposure levels, protective behaviors, or biological factors (such as increased melanin protection) across racial groups. These differences may be influenced by cultural practices, differential access to sun protection resources, varying outdoor activity patterns, or differences in sun safety awareness and education. The association between “no sunburn” responses and climate change harm perceptions suggests that personal experiences with environmental exposures may influence broader environmental health awareness. Respondents reporting no recent sunburn expressed greater uncertainty about climate change health impacts, potentially indicating a disconnect between immediate personal health practices and broader environmental health consciousness.

Public health education efforts should bridge the gap between personal protective behaviors and broader environmental health awareness, fostering comprehensive understanding of how climate change impacts individual and community health.[24] Targeted communication strategies should address specific knowledge gaps identified across demographic groups, such as emphasizing climate change health impacts for populations expressing uncertainty and promoting culturally relevant sun safety practices, particularly for communities with lower sun exposure awareness.[25] Educational resources should be accessible to diverse populations, including those with limited healthcare access or lower health literacy, using plain language and multiple communication channels.[24] By tailoring messages and interventions to the unique needs and perceptions of different demographic groups, public health initiatives can more effectively promote climate resilience and adaptive health behaviors.

### Limitations

This study has several limitations. The data rely on self-reported responses, which may introduce recall bias if participants do not accurately remember or fully disclose their behaviors, experiences, or health conditions. Social desirability bias may also affect responses to sensitive questions about social determinants of health. The relatively modest sample size limits statistical power for detecting small effect sizes, particularly in stratified analyses. The underrepresentation of Asian and other racial/ethnic minority groups may reduce generalizability to these populations and could introduce selection bias. Cross-sectional survey design precludes causal inference regarding observed associations. Future longitudinal studies with larger, more diverse samples are needed to validate these findings and explore temporal relationships between social determinants, health behaviors, and health outcomes in populations with chronic disease comorbidities.[26]

## CONCLUSION

This study reveals important demographic, clinical, and social patterns among individuals with concurrent cancer and hypertension. Significant sex-based and racial/ethnic disparities in social determinants of health, healthcare information sharing comfort, and climate change perceptions highlight the need for targeted, culturally responsive interventions. Healthcare systems should integrate social determinants screening and support into routine clinical care, particularly for vulnerable populations experiencing food insecurity, transportation barriers, and housing instability. Promoting patient-centered care coordination and developing tailored health education addressing both chronic disease management and environmental health awareness may improve outcomes for this complex patient population. Future research should employ longitudinal designs to examine causal relationships and evaluate the effectiveness of interventions addressing the multifaceted needs of patients with multiple chronic conditions.

## CONTRIBUTION STATEMENT

JY designed the study, contributed to the data analyses, and the writing of the manuscript. HC contributed to data analyses and the writing of the manuscript. All authors read and approved the final version of the manuscript.

## DATA AVAILABILITY

Data from the Health Information National Trends Survey (HINTS 6) are publicly available from the National Cancer Institute at https://hints.cancer.gov/

## REFERENCES

1. Sung, H., et al., Global cancer statistics 2020: GLOBOCAN estimates of incidence and mortality worldwide for 36 cancers in 185 countries. CA: a cancer journal for clinicians, 2021. 71(3): p. 209–249.

2. Mills, K.T., A. Stefanescu, and J. He, The global epidemiology of hypertension. Nature Reviews Nephrology, 2020. 16(4): p. 223–237.

3. Chen, H. and J. Ye, Digital health technology burden and frustration among patients with multimorbidity. medRxiv, 2025: p. 2025.10. 09.25337645.

4. Herrmann, J., et al. Evaluation and management of patients with heart disease and cancer: cardio-oncology. in Mayo Clinic Proceedings. 2014. Elsevier.

5. Chung, R., et al., Hypertensive cardiotoxicity in cancer treatment—systematic analysis of adjunct, conventional chemotherapy, and novel therapies—epidemiology, incidence, and pathophysiology. Journal of Clinical Medicine, 2020. 9(10): p. 3346.

6. Ye, J., et al., Characteristics and Patterns of Retention in Hypertension Care in Primary Care Settings From the Hypertension Treatment in Nigeria Program. JAMA Network Open, 2022. 5(9): p. e2230025–e2230025.

7. Finney Rutten, L.J., et al., Data resource profile: the national cancer institute’s health information national trends survey (HINTS). International journal of epidemiology, 2020. 49(1): p. 17–17j.

8. Nelson, D., et al., The health information national trends survey (HINTS): development, design, and dissemination. Journal of health communication, 2004. 9(5): p. 443–460.

9. Ye, J. and Z. Ren, Examining the impact of sex differences and the COVID-19 pandemic on health and health care: findings from a national cross-sectional study. JAMIA Open, 2022.

10. Team, R.C., R: A language and environment for statistical computing. Vienna, Austria: R Foundation for Statistical Computing; 2014. 2019.

11. Chen, H., et al., Telehealth Utilization and Patient Experiences: The Role of Social Determinants of Health Among Individuals with Hypertension and Diabetes. medRxiv, 2024: p. 2024.08. 01.24311392.

12. Ramlakhan, K.P., M.R. Johnson, and J.W. Roos-Hesselink, Pregnancy and cardiovascular disease. Nature Reviews Cardiology, 2020. 17(11): p. 718–731.

13. Ye, J., Z. Wang, and J. Hai, Social Networking Service, Patient-Generated Health Data, and Population Health Informatics: National Cross-sectional Study of Patterns and Implications of Leveraging Digital Technologies to Support Mental Health and Well-being. Journal of Medical Internet Research, 2022. 24(4): p. e30898.

14. Mosca, L., E. Barrett-Connor, and N. Kass Wenger, Sex/gender differences in cardiovascular disease prevention: what a difference a decade makes. Circulation, 2011. 124(19): p. 2145–2154.

15. Ye, J. and Q. Ma. The effects and patterns among mobile health, social determinants, and physical activity: a nationally representative cross-sectional study. in AMIA Annual Symposium Proceedings. 2021. American Medical Informatics Association.

16. Hill-Briggs, F., et al., Social determinants of health and diabetes: a scientific review. Diabetes care, 2020. 44(1): p. 258.

17. Gómez, C.A., et al., Addressing health equity and social determinants of health through healthy people 2030. Journal of public health management and practice, 2021. 27(Supplement 6): p. S249–S257.

18. Marmot, M., et al., WHO European review of social determinants of health and the health divide. The lancet, 2012. 380(9846): p. 1011–1029.

19. Ye, J., et al., Community-Based Participatory Research and System Dynamics Modeling for Improving Retention in Hypertension Care. JAMA Network Open, 2024. 7(8): p. e2430213–e2430213.

20. Kangovi, S., et al., Evidence-Based Community Health Worker Program Addresses Unmet Social Needs And Generates Positive Return On Investment: A return on investment analysis of a randomized controlled trial of a standardized community health worker program that addresses unmet social needs for disadvantaged individuals. Health Affairs, 2020. 39(2): p. 207–213.

21. Ye, J. and S. Bronstein, Using shared clinical decision support to reduce adverse drug events and improve patient safety. Frontiers in Digital Health, 2025. 7: p. 1703141.

22. Peek, M.E., A. Cargill, and E.S. Huang, Diabetes health disparities. Medical care research and review, 2007. 64(5_suppl): p. 101S–156S.

23. Ye, J., et al., Identifying Contextual Factors and Strategies for Practice Facilitation in Primary Care Quality Improvement Using an Informatics-Driven Model: Framework Development and Mixed Methods Case Study. JMIR Human Factors, 2022. 9(2): p. e32174.

24. Romanello, M., et al., The 2021 report of the Lancet Countdown on health and climate change: code red for a healthy future. The Lancet, 2021. 398(10311): p. 1619–1662.

25. Ebi, K.L., et al., Extreme weather and climate change: population health and health system implications. Annual review of public health, 2021. 42(1): p. 293–315.

26. Zhang, S., et al., Machine learning-based mortality prediction in critically ill patients with hypertension: comparative analysis, fairness, and interpretability. Frontiers in Artificial Intelligence, 2025. 8: p. 1686378.

